# A leap to non-communicable diseases epidemic in Burundi: overall trends of the disproportionate burden

**DOI:** 10.1101/2022.09.18.22280066

**Authors:** David Niyukuri, Joseph Nyandwi, Olivier Kamatari, Canesius Uwizeyimana, Chamy Mikaza, Mediatrice Barengayabo

## Abstract

In developing countries, Noncommunicable diseases (NCDs) are increasing relentlessly. A recent review in East African Community countries showed that much need to be done to improve in prevention of risk factors, monitoring, surveillance, governance, and evaluation of health systems to tackle NCDs. In Burundi, there is no study which has been conducted to give an overview on status of NCDs and their risks factors. We conducted a review on estimations of some NCDs statistics and pooled the data of NCDs and associated risk factors, together with data available on cancer for Burundi. A descriptive analysis was performed, and statistical test was conducted to check distribution differences of the NCDs burden between male and female populations. We built linear model for diabetes and hypertension with same predictors (blood pressure, body-mass index, and cholesterol). Overall, data pooled in different studies showed that almost all the NCDs and associated risk factors have an increasing trend in Burundi among male and female populations. Adult men and women are at high risk of developing diabetes and hypertension. Although both female and male populations have an increasing trend of obesity, women, girls between 5 and 10 years, and adolescent girls are carrying the highest burden compared to their male counterpart with same age categories. The data showed that obesity has been sharply increasing among children and adolescents in the last two decades. Cancer is also increasing with a lot of new types being diagnosed. We conclude that, NCDs are of a lot of concerns in male and female populations in Burundi. The prevalence of obesity among younger children and adolescent is alarming. There is a need of actions to be done in order to be able to prevent and manage NCDs, but interventions targeting children, adolescents, and women should be put in place urgently.

## Introduction

According to the World Health organization (WHO), Noncommunicable diseases (NCDs), also known as chronic diseases tend to be of long duration and are the result of a combination of genetic, physiological, environmental, and behavioural factors[1]. The main types of NCD are cardiovascular diseases, cancers, chronic respiratory diseases, and diabetes. Estimates from WHO show that, at global level, NCDs kill 41 million people each year, equivalent to 71% of all deaths globally, and 77% of all NCD deaths are in low- and middle-income countries[1]. Cardiovascular diseases claim 17.9 million lives annually worldwide, followed by cancers (9.3 million), respiratory diseases (4.1 million), and diabetes (1.5 million). According to global burden of disease study in 2017, Sub-Saharan Africa has seen a rapid increase of NCDs burden from 19% to 30% of the total diseases burden, between 1990 and 2017[2]. In a recent review of NCDs in the East Africa Community which includes Burundi, it was reported that cardiovascular diseases caused 13% of all NCDs related deaths, followed by malignant neoplasms (5.9%), respiratory diseases (2.1%), and diabetes mellitus (1.9%) according to data reported between 2000-2016[3].

From Global Cancer Statistics 2020 (GLOBOCAN 2020) report, cancer has reached epidemic proportions worldwide, it claimed almost 10 million individuals, globally[4]. This is more than HIV/AIDS, tuberculosis and malaria combined. As all types of NCDs, most deaths due to cancer occur in people living in low- and middle-income countries (LMIC). In Burundi, the International Agency for Research on Cancer’s (IARC) GLOBOCAN database has showed that in 2020 there were 7929 new cancer cases (3288 in men and 4641 in women) and 5701 cancer deaths (2413 in men and 3288 in women)[5]. According to Bujumbura Pathology Center (BUJAPATH) report, women are more affected with high burden of cervical and breast cancer, and many men are being diagnosed with prostate cancer in Burundi [6].

The current trends of NCDs combined to the burden of infectious diseases show that, it is less likely to achieve the 4^th^ target of the 3^rd^ Sustainable Development Goal (SDG) dedicated to reducing by one third premature mortality from noncommunicable diseases through prevention and treatment, and promote mental health and well-being by 2030[7]. In the mentioned review of NCDs in East Africa, the results of the East African NCD Alliance Benchmark Survey (2017) showed that Burundi was lagging behind compared to others in terms of Governance, Prevention and reduction of risk factors, Health systems readiness, Monitoring, surveillance and evaluation[3].

It is well known that lifestyle behaviours and individuals’ metabolism are the two main types of risk factors. The so called, modifiable behaviours associated to lifestyle such as tobacco use, physical inactivity, unhealthy diet, and the harmful use of alcohol, all increase the risk of NCDs. Metabolic risk factors such as raised blood pressure, overweight/obesity, hyperglycemia (high blood glucose levels), and hyperlipidemia (high levels of fat in the blood) increase the likelihood of having NCDs and dying from it. Most of the time, the leading metabolic risk factor for deaths is elevated blood pressure, followed by overweight and obesity and raised blood glucose[8–10]. Therefore, as long-term diseases with high cost on health systems and families, NCDs need more attention especially for countries with low-income and high proportions of young population such Burundi.

In the EAC review [3], authors reported that in the EAC there is modest progress in governance, prevention of risk factors, monitoring, surveillance, and evaluation of health systems can be observed. Many policies exist on paper, implementation and healthcare are weak and there are large regional and subnational differences. Thus, there is a need to know the temporal trends of NCDs, and their risk factors in Burundi, and what are the magnitudes of different risk factors on NCDs prevalence in Burundi might be. This will help to mobilize national policymakers, non-governmental organizations (NGOs) and other stakeholders to ensure future NCD policies and implementation improvements. For example, Asbestos can be a deadly health hazard such as asbestosis and lung cancer. In Burundi, the chemical substances contained in asbestos-based roofing pose a danger to human health. Inhalation of its fibers can lead to various serious lung diseases, including asbestosis and cancer[11]. However, houses with such roofing material can be seen in many cities in Burundi. According to Worldwide Asbestos supply and Trends report, from 1900 through 2003, in 1984 Burundi has imported 125 metric tons of asbestos roofing [12]. According to DHS analytical study which explored the effects of improved housing conditions on malaria infection in children in Sub-Saharan Africa conducted in 2016, it shows that asbestos/slate roofing sheets are still used in Burundi (Data for Burundi were collected in 2020) [13].

In this study we focus on NCDs conditions and associated risk factors for which data is available in the literature for Burundi. The main objective is to understand the over all trends and distributions differences in the general population, among female and male population, children and adolescents.

## Materials and Methods

This study is a kind of type of NCDs estimates published in different research papers and use data available in public domain, and also from non-published reports from health institutions. Thus, we pooled data from different sources, and analyzed them using Excel, and build basic statistical models for diabetes and hypertension using R[14].

### Data collection

Most of the data we used were gathered from the estimates computed by the NCD Risk Factor Collaboration (NCD-RisC)[15]. The latter is a network of health scientists around the World that provides rigorous and timely data on major risk factors for non-communicable diseases for all of the World’s countries. From the NCD-RisC platform, we downloaded from the platform published data for hypertension, diabetes, cholesterol, blood pressure, and body-mass index [8–10,16–19]. We pooled those annual estimates together for descriptive analysis, and predict future trends based on estimated NCDs risk factors. We also used available data from GLOBOCAN [4] and Bujumbura Pathology Center (BUJAPATH), a cancer screening and diagnosis center located in Bujumbura. Due to the lack of cancer registry in Burundi, only data of 2020 was available in the GLOBOCAN database.

### Data analysis and statistical modelling

Most of the data we used were gathered from the estimates computed by the NCD Risk Factor Collaboration (NCD-RisC). We conducted a descriptive analysis, summarize with means and standard deviations of different factors among male and female populations, and visualized the temporal trends of NCDs burden in Burundi. We also conducted a t-Test of two sample assuming unequal variances to see if the distributions of risk factors were the same in men and women[20].

We built two linear models [21,22], for age-standardized prevalence of hypertension, and diabetes prevalence among adults. The following equation (1) gives the mathematical expression of the statistical linear model, where parameters β give information on the effects of predictors (*x*) to the response variable (*y*), and ε the associated error. In our model, age-standardized prevalence of hypertension, and diabetes prevalence among adults are both considered as response variables (*y*). And the predictor variables (*x*) for both the prevalence of hypertension and diabetes were mean age-standardized BMI, mean total cholesterol, age-standardized mean systolic blood pressure (mmHg), and age-standardized mean diastolic blood pressure (mmHg).

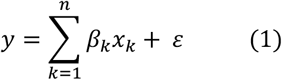

Those predictors are known to be risk factors of those two diseases, and yearly data was available. These two linear models will show the magnitude effects of each predictor variables on each disease. Due to differences in time periods of available data, we considered the shortest period (1990-2014) to make sure we get data for all variables in our models. Prior to modelling, we also conducted a correlation analysis between predictors of diabetes, and hypertension since it is known that these two diseases are somehow interconnected [23,24]. After normality test using Shapiro test, we used Pearson method to conduct the correlation analysis [25,26].

## Results

### Descriptive analysis

The results of the descriptive analysis of hypertension, diabetes, cancer, and their risk factors, show the temporal trends of these diseases and their factors in Burundi in the last decades. We also presented results on the distributions of cases between male and female populations.

### Hypertension

The age-standardized prevalence of hypertension for men and women between 1990 and 2019 shows a decreasing pattern in the last decade as we can see at Figure 1. However, we can see that there is a difference between the two populations. Women with hypertension are decreasing a low speed compared to men, and that difference has becoming bigger with time.

**Figure 1.**
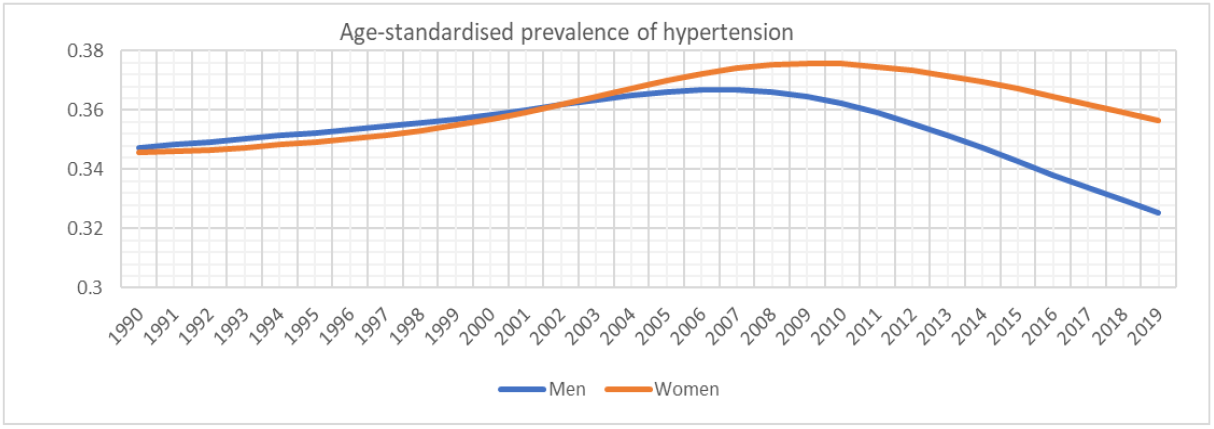
Age-standardized prevalence of hypertension in adult population between 1990 and 2019.

As we can see in Table 1, the mean value (and standard deviation) of age-standardized prevalence of hypertension for men and women was 0.353 (0.011) and 0.361 (0.010), respectively. The t-Test between the two population shows that the two populations had different distribution (p=0.005). From Table 1, we can see that on average, only around 20% of hypertension cases were diagnosed among all hypertension cases. The mean value (and standard deviation) of age-standardised proportion of diagnosed hypertension among all hypertension for men and women was 0.2286 (0.0504) and 0.3476 (0.0620), respectively. The mean value (and standard deviation) of age-standardised proportion of treated hypertension among all hypertension for men and women was 0.1183 (0.0411) and 0.1877 (0.0568), respectively. And the mean value (and standard deviation) of age-standardised proportion of controlled hypertension among all hypertension for men and women was 0.0368 (0.0224) and 0.0539 (0.0325), respectively. The t-Test for the 3 measurements between men and women shows that the two populations had different distributions with p<0.0001) for age-standardised proportion of diagnosed hypertension, age-standardised proportion of treated hypertension, and age-standardised proportion of controlled hypertension.

**Table 1.** Summary of temporal trends of NCDs burden in Burundi.

| Indicator | Period | Male |  | Female |  | p.Value |
| --- | --- | --- | --- | --- | --- | --- |
|  |  | Mean/SD | OR (95%CI) | Mean/SD | OR (95%CI) |  |
| Age-standardised prevalence of hypertension | 1990-2019 | 0.3535 / 0.0111 | 0.0041 | 0.36145/0.0104 | 0.0039 | 0.0054 |
| Age-standardised proportion of diagnosed hypertension among all hypertension | 1990-2019 | 0.2287/0.0504 | 0.0188 | 0.3476/0.062 | 0.0232 | 4.3459E-11 |
| Age-standardised proportion of treated hypertension among all hypertension | 1990-2019 | 0.1184/0.0412 | 0.0154 | 0.1877/0.0569 | 0.0212 | 1.5277E-06 |
| Age-standardised proportion of controlled hypertension among all hypertension | 1990-2019 | 0.0368/0.0224 | 0.0084 | 0.0539/0.0326 | 0.0122 | 0.0122 |
| Age-standardised diabetes prevalence | 1980-2014 | 0.0236/0.0099 | 0.0034 | 0.0262/0.0078 | 0.0027 | 0.2237 |
| Mean total cholesterol (mmol/L) | 1980-2018 | 3.7162/0.0332 | 0.0107 | 3.9172/0.1052 | 0.0341 | 7.69E-15 |
| Age-standardized mean systolic blood pressure (mmHg) | 1975-2015 | 126.8130/2.1132 | 0.6670 | 123.6309/3.1091 | 0.9814 | 7.99E-07 |
| Age-standardised mean diastolic blood pressure (mmHg) | 1975-2015 | 75.5575/1.5039 | 0.4747 | 76.3445/2.2031 | 0.6954 | 0.063 |
| Age-standardised prevalence of raised blood pressure | 1975-2015 | 0.2289/0.0274 | 0.0087 | 0.2574/0.0332 | 0.0105 | 5.86E-05 |
| Mean BMI | 1975-2016 | 20.4302/1.0290 | 0.3206 | 19.844/1.1141 | 0.3472 | 0.0142 |
| Prevalence of BMI≥30 kg/m<sup>2</sup> (obesity) | 1975-2015 | 0.0084/0.0058 | 0.0018 | 0.0376/0.0236 | 0.0073 | 6.02E-10 |
| Prevalence of BMI≥35 kg/m<sup>2</sup> (severe obesity) | 1975-2015 | 0.0006/0.0007 | 0.0002 | 0.0069/0.0065 | 0.0020 | 1.75E-07 |
| Prevalence of BMI ≥40 kg/m<sup>2</sup> (morbid obesity) ( | 1975-2015 | 0.0001/0.0002 | 0.0001 | 0.002/0.0022 | 0.0007 | 3.16E-06 |
| Mean BMI 5-10years | 1985-2019 | 14.0309/0.3889 | 0.1336 | 14.1954/0.0999 | 0.0343 | 0.02026 |
| Mean BMI 11-19years | 1985-2019 | 17.9916/0.3889 | 0.1336 | 19.0709/0.0999 | 0.0343 | 2.21E-18 |
| Prevalence of BMI>2SD (obesity) 5-10years | 1975-2015 | 0.0022/0.0029 | 0.0009 | 0.0058/0.0072 | 0.0022 | 0.0038 |
| Prevalence of BMI>2SD (obesity) 11-19years | 1975-2015 | 0.0015/0.0020 | 0.0006 | 0.0039/0.0048 | 0.0015 | 0.0038 |
| Prevalence of BMI>1SD (overweight)_5-10years | 1975-2015 | 0.0221/0.0201 | 0.0063 | 0.059/0.0425 | 0.0133 | 4.18E-06 |
| Prevalence of BMI>1SD (overweight)_11-19years | 1975-2015 | 0.0172/0.0154 | 0.0048 | 0.0537/0.0375 | 0.0117 | 3.18E-07 |
| Mean BMI (urban) | 1985-2017 | 22.3392/0.6257 | 0.2219 | 22.9738/0.7362 | 0.2610 | 0.0004 |
| Mean BMI (rural) | 1985-2017 | 21.1509/0.0987 | 0.2009 | 20.2505/0.069 | 0.1405 | 2.66E-10 |
| Mean BMI (urban-rural difference) | 1985-2017 | 1.1883/0.0593 | 0.0210 | 2.7234/0.3439 | 0.1220 | 1.29E-23 |

### Diabetes

The age-standardized prevalence of diabetes for men and women between 1980 and 2014 shows an increasing trend in the last 3 decades as we can see at Figure 2. In 1980s, women had high prevalence compared to men. However, that difference between the two populations has been decreasing for years, and it reached a point in 2010s where the prevalence of men became higher compared to women. The age-standardized prevalence of diabetes can be considered relatively close to one another in women and men.

**Figure 2.**
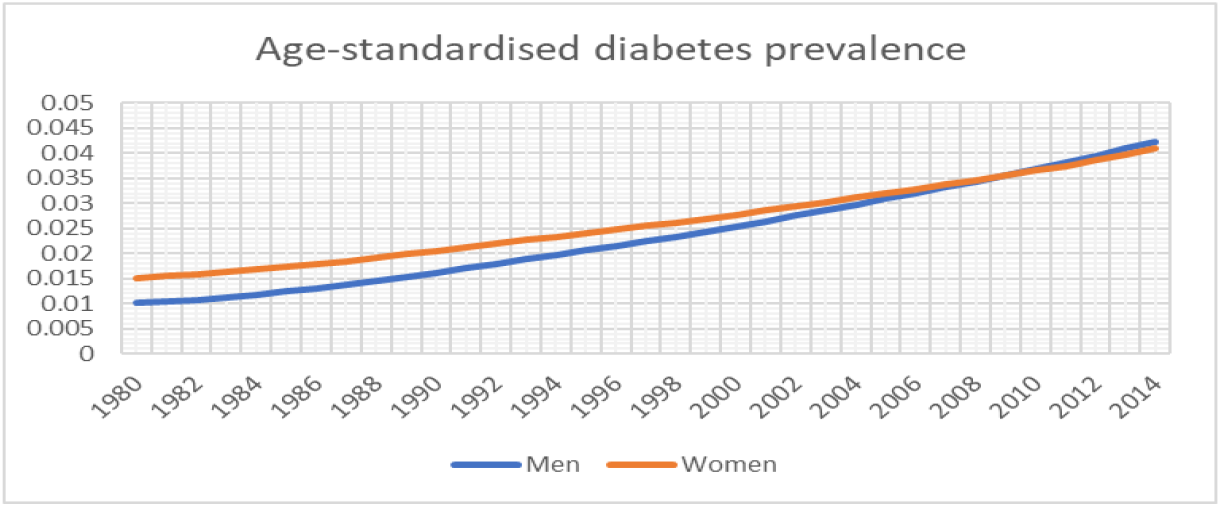
Age-standardized prevalence of diabetes in adult population between 1990 and 2019.

From Table 1, the mean value (and standard deviation) of age-standardized prevalence of diabetes for men and women was 0.0236 (0.0099) and 0.0262 (0.0077), respectively. The t-Test between the two population shows that the two populations had same distribution (p=0.2236).

### Obesity in general population

The overall obesity in the general population, severe obesity, and morbid obesity for men and women between 1975 and 2015 shows increasing trends for both genders as we can see at Figure 3 below. However, women’s obesity prevalence has the fastest increasing trend compared to men. Although they had higher prevalence even in 1980s and 1990s, but it widens much in 2000s.

**Figure 3.**
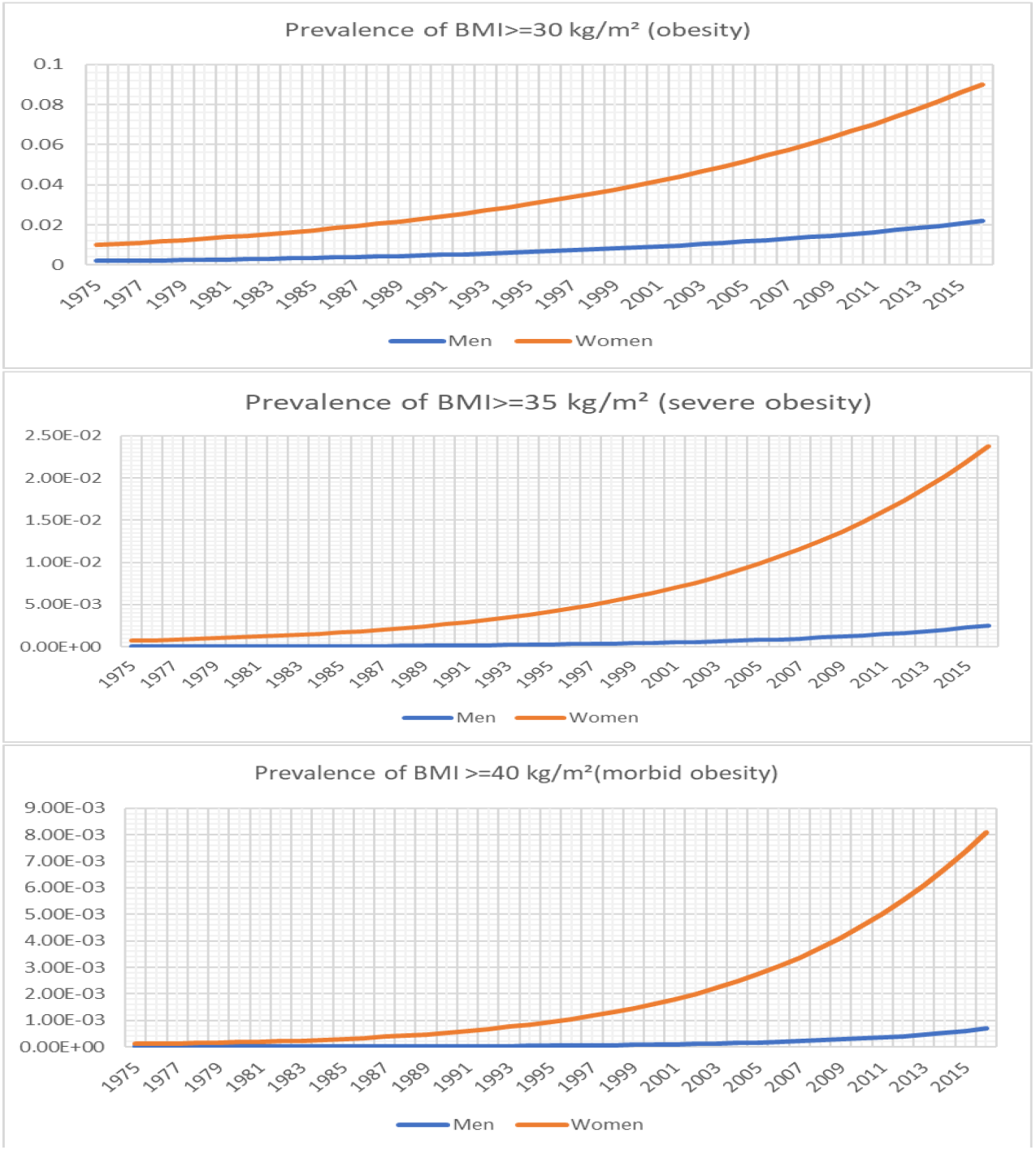
Prevalence of obesity, severe obesity, and morbid obesity (based on BMI) in adult population between 1975 and 2015.

From Table 1, the mean value (and standard deviation) of prevalence of obesity was 0.0084 (0.0058) and 0.0376 (0.0235), for severe obesity it was 0.0006 (0.0006) and 0.0069 (0.0065), and for morbid obesity it was 0.0001 (0.0001) and 0.0019 (0.0022), for men and women, respectively. The t-Test for the 3 measurements between men and women shows that the two populations had different distributions with p<0.0001) for prevalence of obesity, severe obesity, and morbid obesity. In Table 1, we can also see the difference between BMI in urban and rural populations. Between 1985 and 2017, the mean value (and standard deviation) of BMI in urban was 22.3391 (0.6257) and 22.9738 (0.7362) for men and women, respectively. Whereas it was estimated to be 21.1509 (0.0986) and 20.2504 (0.0689) for men and women in rural.

### Obesity in children between 5 and 10 years

When looking at children population, boys, and girls between 5 and 10 years, the prevalence of obesity and overweight have same increasing trends between 1975 and 2019 can be seen at Figure 4. We can also see that, like in the previous result of obesity in the general population, girls’ prevalence has the fastest increasing trend compared to boys. In Table 1, we can see that the mean value (and standard deviation) for obesity was 0.0022 (0.0029) and 0.0059 (0.0071), and for overweight 0.0220 (0.0200) and 0.0589 (0.0425), for boys and girls, respectively. When looking at the BMI measurements in Table 1, for those children between 5 and 10 years from 1985 up to 2019, the mean value (and standard deviation) was 14.0308 (0.3889) and 14.1953 (0.0999), for boys and girls, respectively.

**Figure 4.**
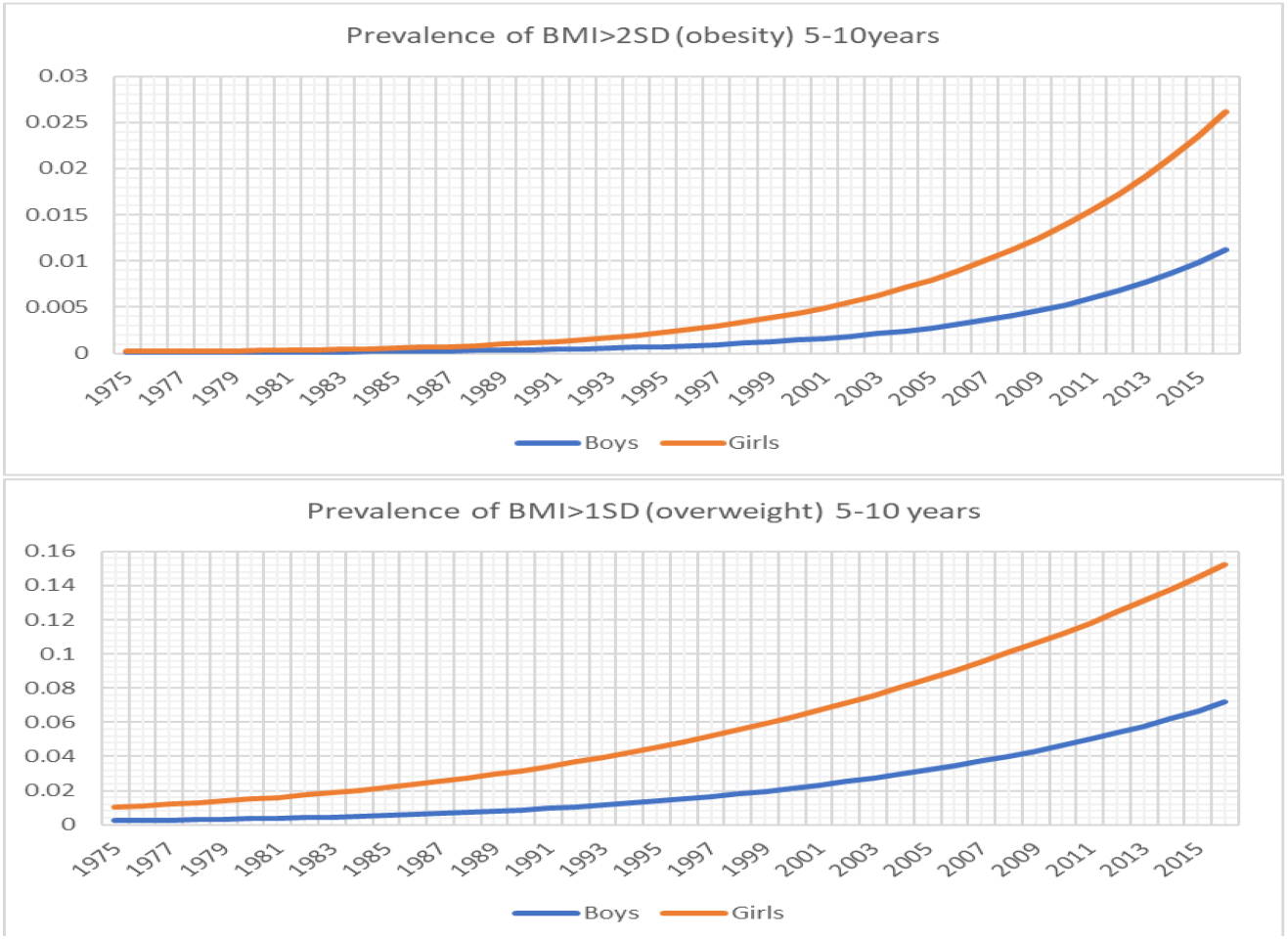
Prevalence of obesity and overweight (based on BMI) among 5-10 years children between 1975 and 2015.

The t-Test for the 2 measurements between boys and girls shows that the two populations had different distributions with p<0.005 for prevalence of obesity, and overweight.

### Obesity in children between 11 and 19 years

When we look at adolescence age groups for boys and girls on Figure 5, we can see same trends as for those between 5 and 10 years. Between 1975 and 2019, Figure 5 shows that girls’ prevalence has the fastest increasing trend compared to boys. In Table 1, we can see that the mean value (and standard deviation) for obesity was 0.00147 (0.0019) and 0.0038 (0.0047), and for overweight 0.0172 (0.0153) and 0.0537 (0.0375), for boys and girls, respectively. For this age category, when looking at the BMI in Table 1, for those children between 11 and 19 years from 1985 up to 2019, the mean value (and standard deviation) was 17.9916 (0.3889) and 19.0708 (0.0998), for boys and girls, respectively.

**Figure 5.**
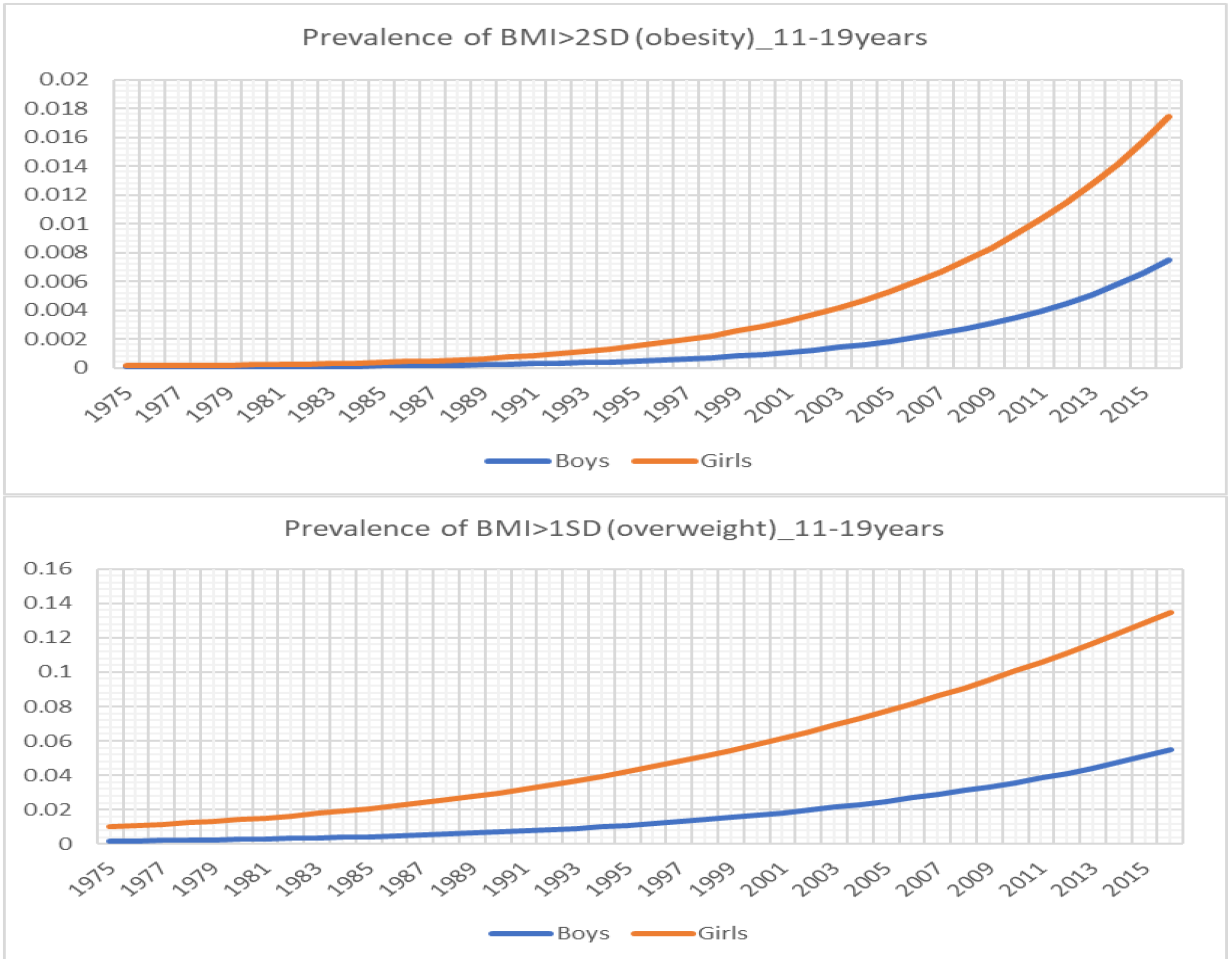
Prevalence of obesity and overweight (based on BMI) among 11-19 years children between 1975 and 2015.

The t-Test for the 2 measurements between boys and girls shows that the two populations had different distributions with p<0.0001 for prevalence of obesity, and overweight.

### Cholesterol

It is well known that high blood pressure (hypertension) and high cholesterol also are linked. As we can see at Figure 6, the level of cholesterol has been increasing. But women have the highest level compared to men. The mean (and standard deviation) of cholesterol level between 1980 and 2018 was 3.7162 (0.0331) and 3.9172 (0.1051) for men and women, respectively. Another important note is that the difference between men and women has been also increasing the gap between the two population and has become much wider. In addition, the t-Test for the cholesterol level between men and women shows that the two populations had different distributions with p<0.0001.

**Figure 6.**
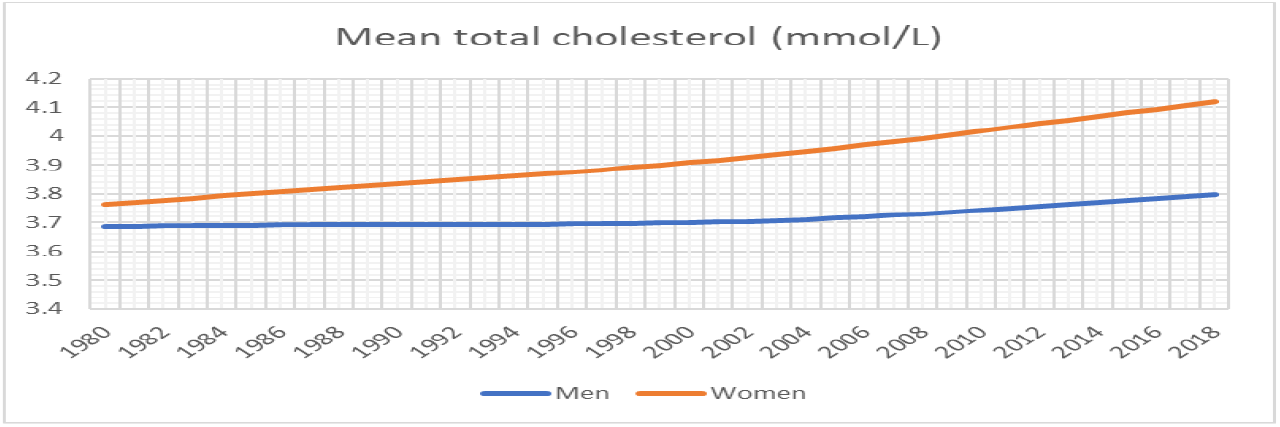
Mean total cholesterol in adult population between 1980 and 2018.

### Blood pressure

High blood pressure known also as hypertension has been increasing in men and women in the last decades (1975-2015) as we can see at Figure 7. Systolic and Diastolic blood pressure have been increasing in men and women. But men have higher mean value of blood pressure for the systolic blood pressure. In addition, the increasing trends between the two populations is different, before 1980s the difference of systolic blood pressure between men and women was huge, but as years come to pass it decreases. Opposite to diastolic blood pressure, women have much higher blood pressure values. Before 1980s men had high blood pressure, but women overtook the trends with high blood pressure values from early 1980s up to 2015. For diastolic blood pressure, the difference between men and women increased.

**Figure 7.**
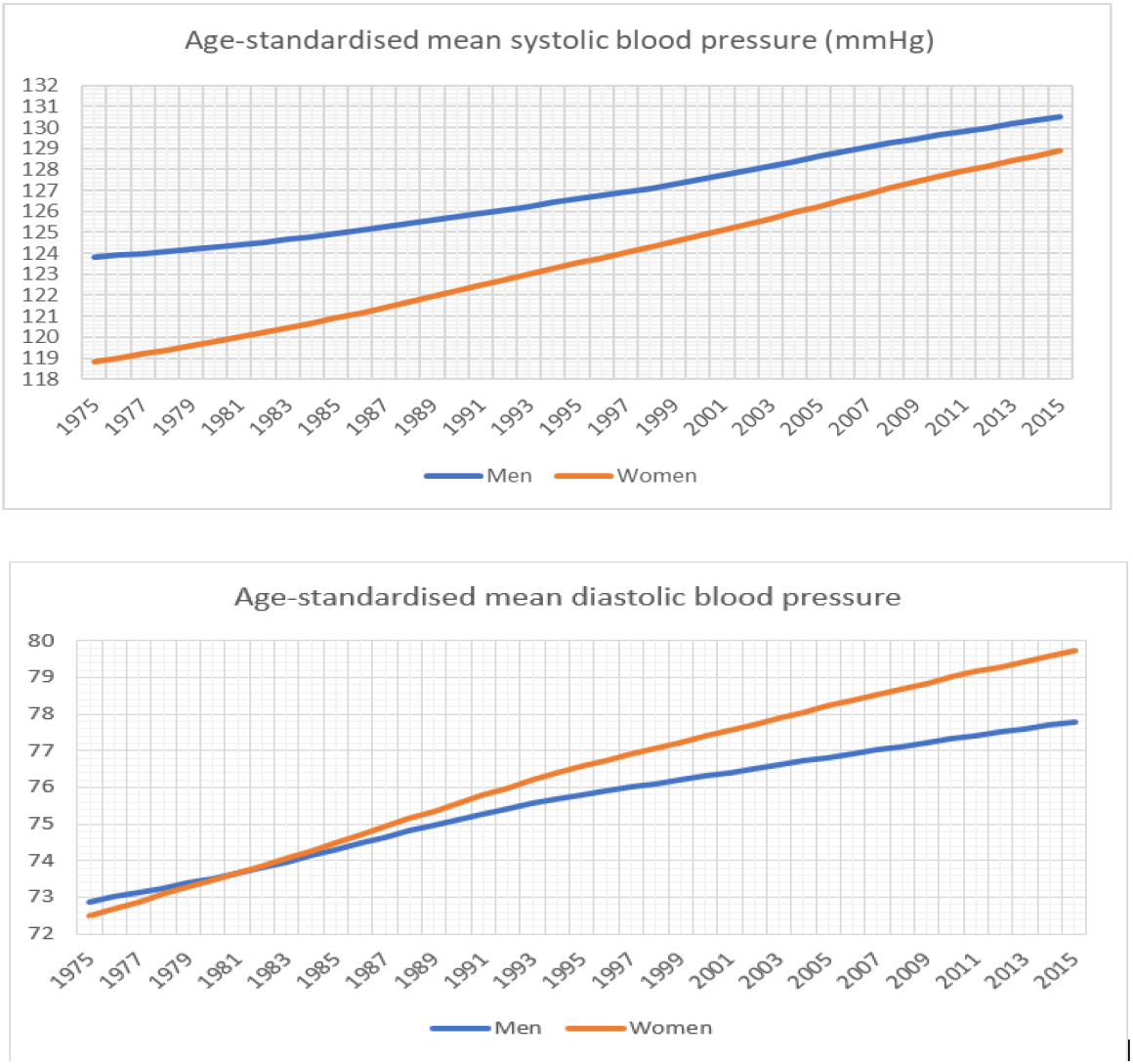
Age-standardized mean blood pressure between 1975 and 2015.

In Table 1, we can see that the mean value (and standard deviation) for the age-standardized mean systolic blood pressure was 126.8130 (2.1131) and 123.6308 (3.1091) for men and women, respectively. But the mean value (and standard deviation) for the age-standardized mean diastolic blood pressure was 75.5575 (1.5038) and 76.3445 (2.2031) for men and women, respectively. The t-Test for the age-standardized mean systolic blood pressure between men and women shows that the two populations had different distributions with p<0.0001. However, for the age-standardized mean diastolic blood pressure the p>0.05 (p=0.0629). Which means that the two distributions of diastolic blood pressure for men and women were the same. Furthermore, we can see in Table 1 that, on average, between 1975 and 2015, the age-standardized prevalence of raised blood pressure was 0.2288 (0.0274) and 0.2573 (0.0331) for men and women, respectively.

### Cancer

At Figure 8, panel A and B show the increasing trends of new cases of cancer for different types of cancer in Burundi. We can see at panel A most prevalent cancer from different parts of human body such as colorectum, stomach, breast, prostate, skin, and head and neck. In reported data of 2019 and 2020 women has lower proportion compared to men as we can see at panel B, but there has been a sharp increase of women patients in 2021.

**Figure 8.**
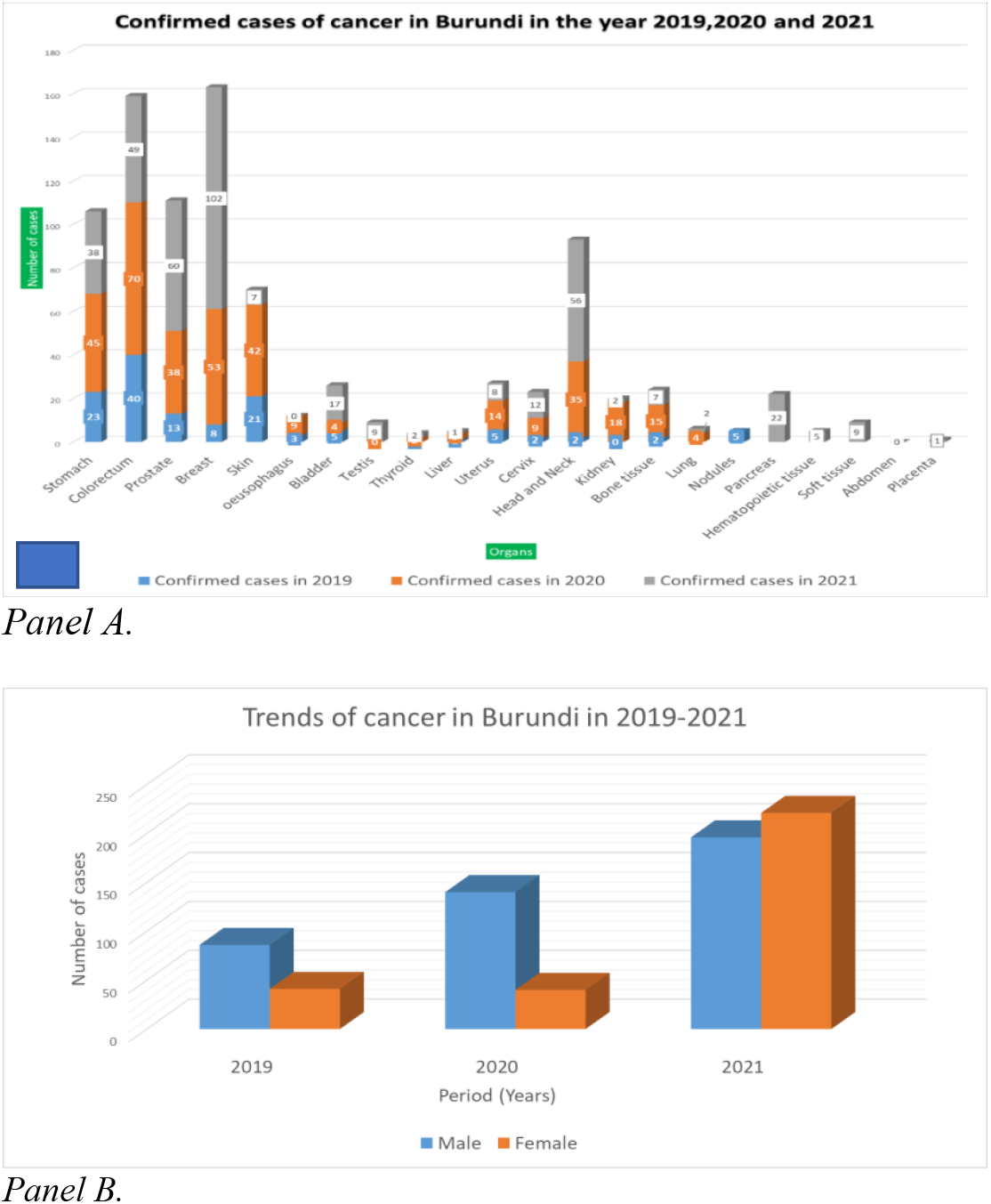
Panel A. Confirmed cancer cases by types and proportion of men and women patients in Burundi between 2019-2021 Panel B. Trends of confirmed cancer cases among men and women between 2019 and 2021.

In supplement information S1, the Figure shows the trends of cancer types by positivity rates in the last 3 years. We can see that for some types of cancer with more than 10 cases such as colorectal, stomach, breast, we had always more than 40% of positivity rate at least once in the 3 years. When looking to age categories of cases, we can see that all categories are affected by the disease. But much burden can be observed for 41-60 years, and beyond for both genders. However, for adult population between 21 and 40 years, women carry a disproportionate burden of the disease as we can see it in supplement information S2.

### Models’ outputs

The normality test of all predictors and the response variables showed that except for women BMI and men systolic blood pressure, the approximate p-value was greater than 0.05, which means that their distributions were not significantly different from normal distributions. The correlations analysis with the Pearson method (in supplement information S3) showed much correlation between predictors (BMI, cholesterol, and blood pressure), and between them and response variables (diabetes and hypertension).

The estimates which show the effect of the predictors (BMI, blood pressure, and cholesterol) to the hypertension and diabetes models had significant effects for both men and women as we can see in Table 2. For diabetes in men and women, the BMI and cholesterol had same increase effects but with different magnitude. Parameter estimates associated to both predictors were positive with high magnitude for men. Thus, an increase of BMI or cholesterol was associated to an increase of diabetes cases for men and women. For blood pressure, we can see that diastolic blood pressure had negative parameter estimate for men (opposite to systolic blood pressure which had positive parameter estimate) whereas it had positive parameter estimate for women (oppositive to systolic blood pressure which had negative parameter estimate for women). For hypertension in men and women, the BMI and cholesterol had same effects but with different magnitude. Parameter estimates associated to both predictors were negative with high magnitude for women. For blood pressure, we can see that except diastolic blood pressure for women other risk factors had also positive parameter estimates.

**Table 2.** Estimates of predictors effects on diabetes and hypertension among men and women populations in Burundi. Significant codes: 0 ‘***’ 0.001 ‘**’ 0.01 ‘*’ 0.05 ‘.’ 0.1 ‘ ‘ 1

|  | Estimate | Std | t-value | Pr(>\|t\|) | R-square |
| --- | --- | --- | --- | --- | --- |
| **Diabetes (men)** |  |  |  |  |  |
| Intercept | -0.3414190 | 0.1101033 | -3.101 | 0.005633 ** | 0.9998 |
| BMI | 0.0209817 | 0.0046334 | 4.528 | 0.000205 *** |  |
| Cholesterol | 0.1372551 | 0.0150603 | 9.114 | 1.47e-08 *** |  |
| Systolic blood pressure | 0.0012488 | 0.0003177 | 3.931 | 0.000826 *** |  |
| Diastolic blood pressure | -0.0096943 | 0.0033417 | -2.901 | 0.008834 ** |  |
| **Diabetes (women)** |  |  |  |  |  |
| Intercept | -0.2127959 | 0.0239368 | -8.890 | 2.20e-08 *** | 0.9999 |
| BMI | 0.0053690 | 0.0012113 | 4.432 | 0.000256 *** |  |
| Cholesterol | 0.0721665 | 0.0027944 | 25.825 | < 2e-16 *** |  |
| Systolic blood pressure | -0.0016336 | 0.0002998 | -5.450 | 2.47e-05 *** |  |
| Diastolic blood pressure | 0.0006905 | 0.0006219 | 1.110 | 0.280042 |  |
| **Hypertension (men)** |  |  |  |  |  |
| Intercept | -1.143868 | 0.670756 | -1.705 | 0.103615 | 0.09906 |
| BMI | -0.128774 | 0.028227 | -4.562 | 0.000189 *** |  |
| Cholesterol | -1.238517 | 0.091748 | -13.499 | 1.65e-11 *** |  |
| Systolic blood pressure | 0.040623 | 0.001935 | 20.992 | 4.28e-15 *** |  |
| Diastolic blood pressure | 0.047214 | 0.020358 | 2.319 | 0.031080 * |  |
| **Hypertension (women)** |  |  |  |  |  |
| Intercept | -0.967745 | 0.150078 | -6.448 | 2.74e-06 *** | 0.9990 |
| BMI | -0.018933 | 0.007595 | -2.493 | 0.0216 * |  |
| Cholesterol | -0.726876 | 0.017520 | -41.487 | < 2e-16 *** |  |
| Systolic blood pressure | 0.074001 | 0.001879 | 39.374 | < 2e-16 *** |  |
| Diastolic blood pressure | -0.060551 | 0.003899 | -15.528 | 1.27e-12 *** |  |

When we visualize the response variables (diabetes and hypertension) together with the predictors (blood pressure, BMI, and cholesterol), at Figure 9 for diabetes, we can see that the response variable (diabetes prevalence) has an exponential with all predictors (BMI, blood pressure, and cholesterol). This means that, diabetes prevalence increases as the predictors increase. However, for Figure 10 for hypertension, we can see that the response variable (hypertension prevalence) has a quadratic relation with all predictors (BMI, blood pressure, and cholesterol). From that quadratic form, we can see that as the predictors increase the hypertension reaches a maximum prevalence and starts decreasing as the predictors increase beyond the critical value. In other words, beyond critical values of predictors, hypertension prevalence decreases.

**Figure 9.**
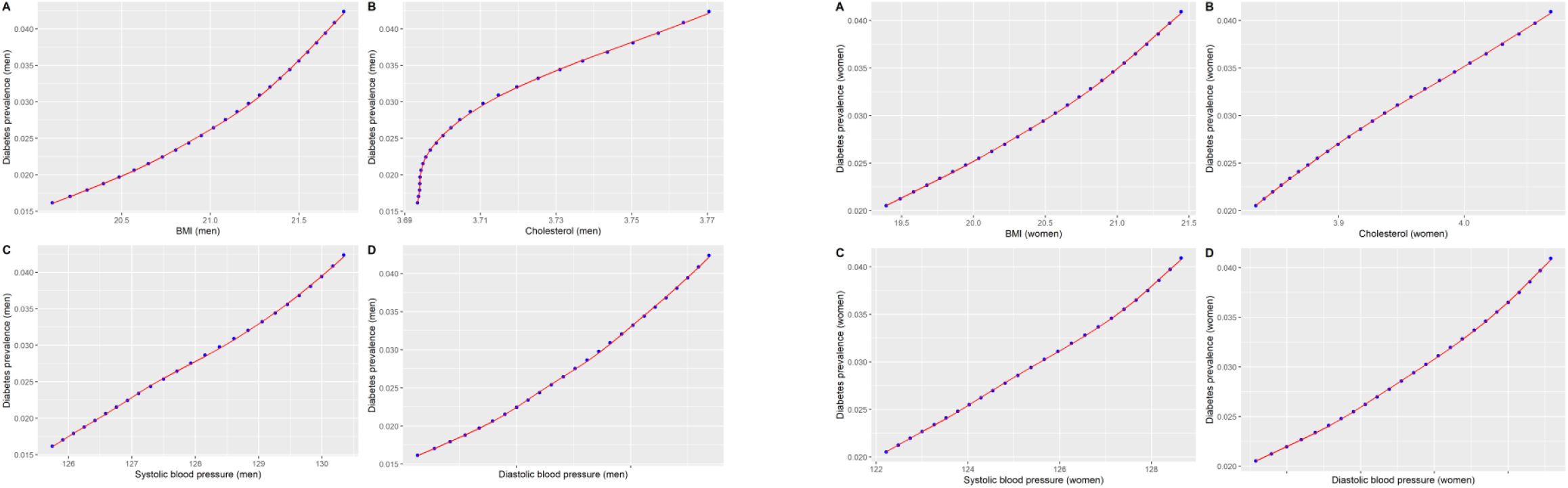
Predictors (BMI, blood pressure, and cholesterol) of diabetes for men and women in Burundi

**Figure 10.**
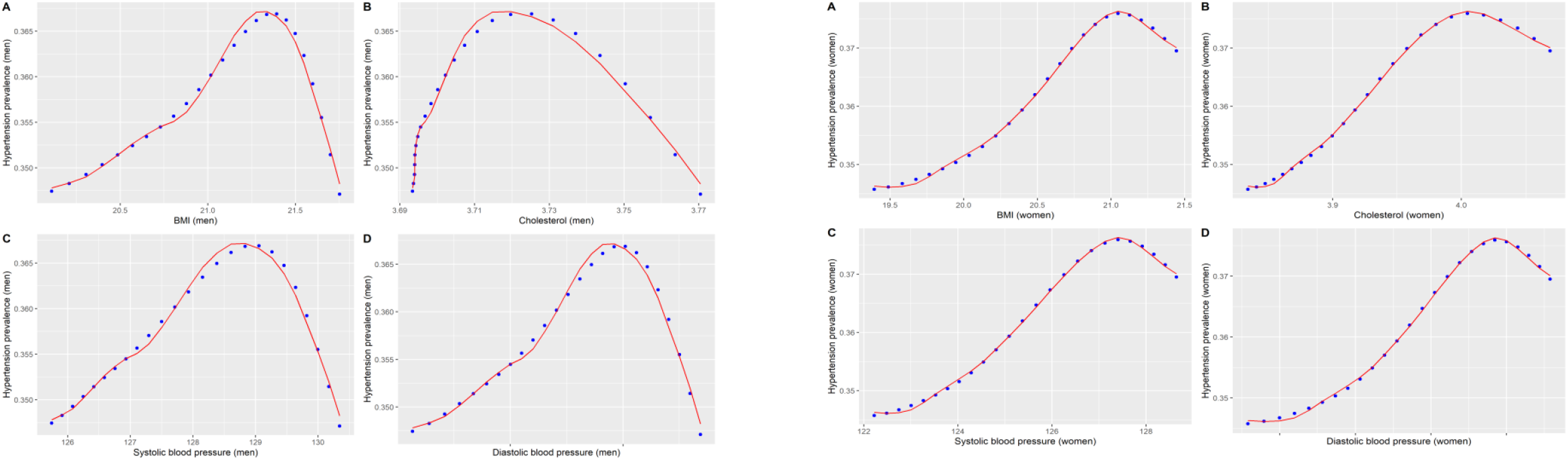
Predictors (BMI, blood pressure, and cholesterol) of hypertension for men and women in Burundi

## Discussion

Overall, data pooled in different studies showed that almost all the NCDs have increasing trends in Burundi, among men and women, except hypertension. The latter is decreasing after many years being high. The difference of burden distributions between female and male populations is seen for almost all the NCDs and associated risk factors, with women having highest prevalence. The diabetes trends are increasing for both populations, but men are having a slightly higher prevalence than women. But the distributions of the prevalence between those two populations are similar. A deep analysis, shows that, although both genders have a very fast increasing trends of obesity, women, girls between 5 and 10 years, and adolescent girls are carrying the highest and fastest increasing burden of obesity compared to their oppositive gender of same age categories. The high obesity prevalence among children and adolescents put them at high risk of acquiring diabetes and hypertension in near future, and early death for adult persons. For cancer, Figures 8 shows the increasing trends of new cases of cancer for different types of cancer in Burundi, and women have high proportion compared to men. This increase may be explained by the availability of diagnosis capacity from different initiatives in Burundi, and mobilization campaigns for women for breast cancer [27].

The models outputs showed that, for diabetes, as the BMI, blood pressure and cholesterol increase, there is an increase of diabetes prevalence. But for hypertension, the relationship has a quadratic form with the BMI, blood pressure, and cholesterol showed that hypertension prevalence reaches a maximum value and starts decreasing as the predictors increase beyond the critical values. Which means that, we cannot truly predict the hypertension prevalence increase with the given predictors. It has to consider other individual level variables such as psycho-social factors among others.

In some studies, conducted in the East Africa and sub-Saharan Africa in the last few years, they showed that NCDs represent a largely ‘silent’ epidemic in Sub-Saharan region. It has a rapid increase in NCDs with a distribution in proportion of 19% to 30% between 1990 and 2007 [3]. The region has a disproportionate burden of both infectious and chronic diseases compared with other world regions. According to the WHO disease burden and mortality estimates, the proportion of all deaths in the WHO Africa region that are attributable to NCDs increased from 22.8% (2.2 million) in the year 2000 to 34.2% (3.0 million) in the year 2016 [3]. In 2013, apart from Burundi, all countries of the EAC have already set up some mechanism to undertake periodic surveillance of NCDs (monitoring, surveillance, and evaluation) and their risk factors according to the report of the WHO non-communicable diseases (NCD) progress monitor for East African community (2017/2020).

This study was a review of existing estimates from the literature, and cancer data from GLOBOCAN [4] and BUJAPATH. For cancer, different initiatives are filling the gaps in different aspects of preventing, treating, and managing the disease [27], although much efforts still needed to alleviate the burden as discussed by Manirakiza et al [28]. However, there is a lack of systematic data on diagnosis of diabetes and hypertensions, and limited diagnostics for cancer screening. In Burundi, there is no systematic Surveillance of risk factors for noncommunicable diseases as per the WHO STEPwise approach [3,29,30], which is implemented in other East African countries. Due to limited data, the results in this study do not give a full picture of the epidemiological situation of NCDs in Burundi. However, the estimates from the consolidated studies conducted in countries with demographics, economy, and health systems similar to Burundi, gave a quick overview of the situation of NCDs in Burundi.

## Conclusion

The results showed that the trends of NCDs and associated risk factors in Burundi are of a lot of concerns in men and women populations. Adult men and women are at high risk of developing diabetes and hypertension. The prevalence of obesity among younger children and adolescent is alarming. This quick scan review study shows that there is an urgent need to discourage unhealthy diets, exposure to tobacco smoke, or other cancerogenic substances and chemical or the harmful use of alcohol. Physical activities play an important protective role for hypertension and diabetes [23], thus, people have to take time for physical activities. Physical inactivity, and health lifestyle should be encouraged by parents, and employers. We have seen that for only the 3 years (2019-2021), many different types of cancer were identified in Burundi. Mainly, cancer of colorectum, stomach, breast, prostate, skin, and head and neck. Thus, sensitization campaigns should extend the target public besides women, and diagnostics technology should be deployed and followed by treatment and other cancer care interventions.

Furthermore, most NCDs diseases can also be attributed to rapid unplanned urbanization, globalization of unhealthy lifestyles and population ageing. Unhealthy diets and a lack of physical activity may raise blood pressure, increase blood glucose, elevate blood lipids and obesity. At individual level, lifestyle is a predominant factor which can help to fight effectively NCDs. Interventions targeting children and women should be put in place. Moreover, institutional lead interventions in schools, workplaces, should be also encouraged.

NCDs threaten progress towards the 2030 Agenda for Sustainable Development, which includes a target of reducing premature deaths from NCDs by one-third by 2030. They have strong negative impact on socio-economy status of countries. In a report by The Economist, authors highlighted the growing burden of NCDs in low- and lower-middle-income countries [31]. From different studies, and reports from WHO, the burden of NCDs in developing countries is increasing relentlessly. Unfortunately, existing healthcare systems in sub-Saharan Africa are not ready to manage these conditions which cost much to families and Governments. The rapid rise in NCDs will likely impede poverty reduction initiatives in low-income countries, particularly by increasing household costs associated with long time health care due to NCDs conditions. Vulnerable and socially disadvantaged people get sicker and die sooner than people of higher social positions, especially because they are at greater risk of being exposed to harmful products, such as tobacco, or unhealthy dietary practices, and have limited access to health services. In low-resource settings, health-care costs for NCDs quickly drain household resources. The exorbitant costs of NCDs, including treatment, which is often lengthy and expensive, combined with loss of income, force millions of people into poverty annually and stifle development.

The lack of availability of cancer therapy services is due to poor implementation of the national policy against cancer. This has resulted in the lack of cancer register, no existence of cancer control infrastructures, poor early detection and diagnosis and a lack of properly trained human resources. The effects of the main problems mentioned above, as well as the lack of awareness of the population on cancer risk factors and the demographic and lifestyle changes, have increased the prevalence of cancer and related mortality rates. The increased prevalence of cancer within Burundi’s population also affects the population’s economy, due to rising health care costs which engenders poverty. Therefore, an important way to control NCDs is to focus on reducing the risk factors associated with these diseases. Low-cost solutions exist for governments and other stakeholders to reduce the common modifiable risk factors. Monitoring progress and trends of NCDs and their risk is important for guiding policy and priorities. To lessen the impact of NCDs on individuals and society, a comprehensive approach is needed requiring all sectors, including health, finance, transport, education, agriculture, planning and others, to collaborate to reduce the risks associated with NCDs, and to promote interventions to prevent and control them. Investing in better management of NCDs is critical. Management of NCDs includes detecting, screening, and treating these diseases, and providing access to palliative care for people in need. High impact essential NCD interventions can be delivered through a primary health care approach to strengthen early detection and timely treatment. Evidence shows such interventions are excellent economic investments because, if provided early to patients, they can reduce the need for more expensive treatment.

## Data Availability

https://ncdrisc.org/.
Published 2022

## Declarations

The findings and conclusions in this article are those of the authors and do not necessarily represent the official position, decisions, policy, or views of these institutions.

## Authors contributions

DN, JN, and MB conceptualized the study. DN, CU, and CM collected and collated the data. OK and DN cleaned the data. OK, DN, CU conducted the analysis. DN wrote code for modelling and wrote the first draft of the manuscript. MB, JN revised extensively the first draft of the manuscript. All authors wrote and reviewed the manuscript.

## Ethical statement

We used data available in public domain.

## Acknowledgments

Authors thank the Doctoral School of the University of Burundi for supporting this study.

## Supplement Information

### S1: Cancer Positivity rates trends by types in Burundi between 2019 and 2021

**Figure.**
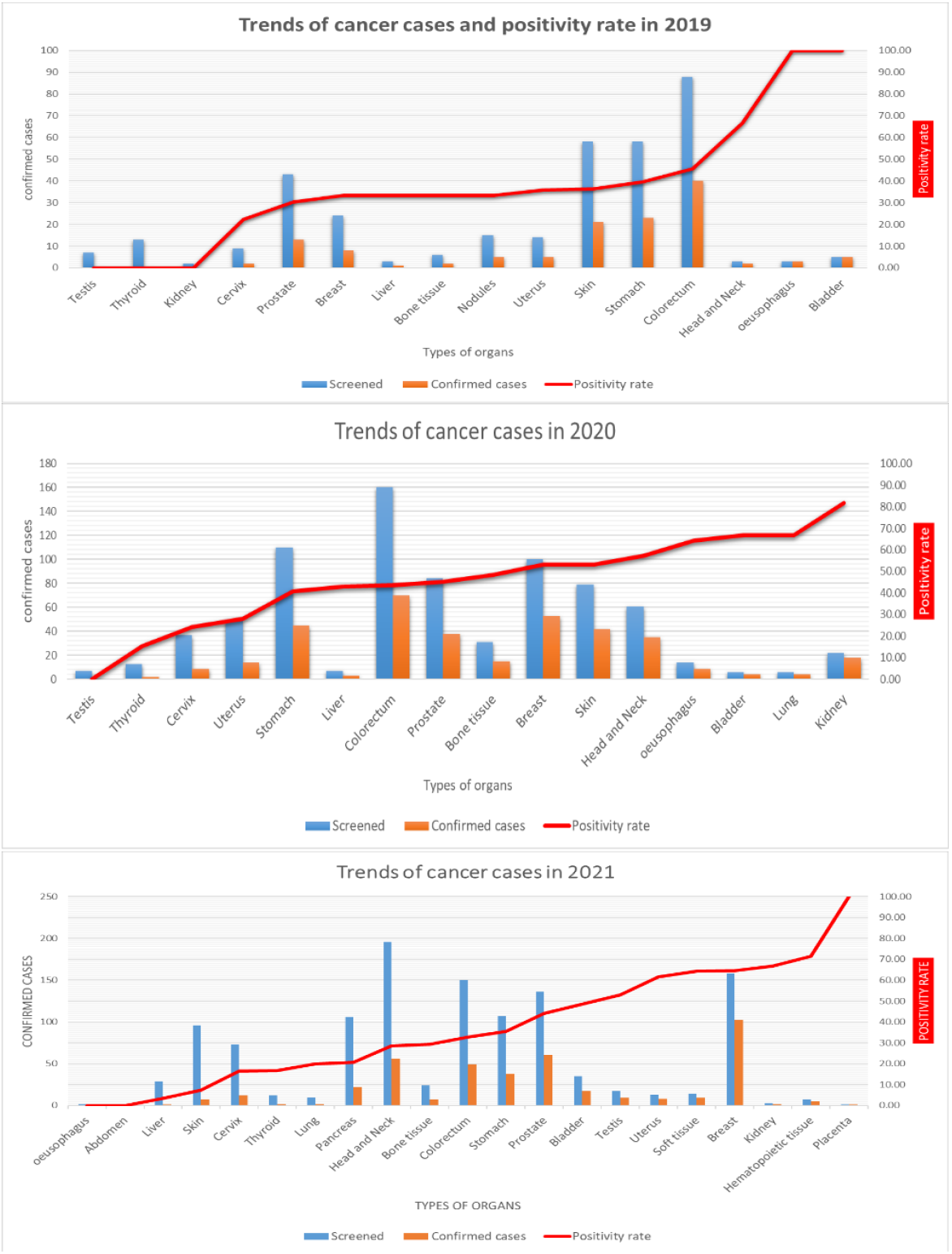
Positivity rates trends of cancer cases by types between 2019-2021.

### S2: Age groups of cancer patients in Burundi between 2019 and 2021

**Figure.**
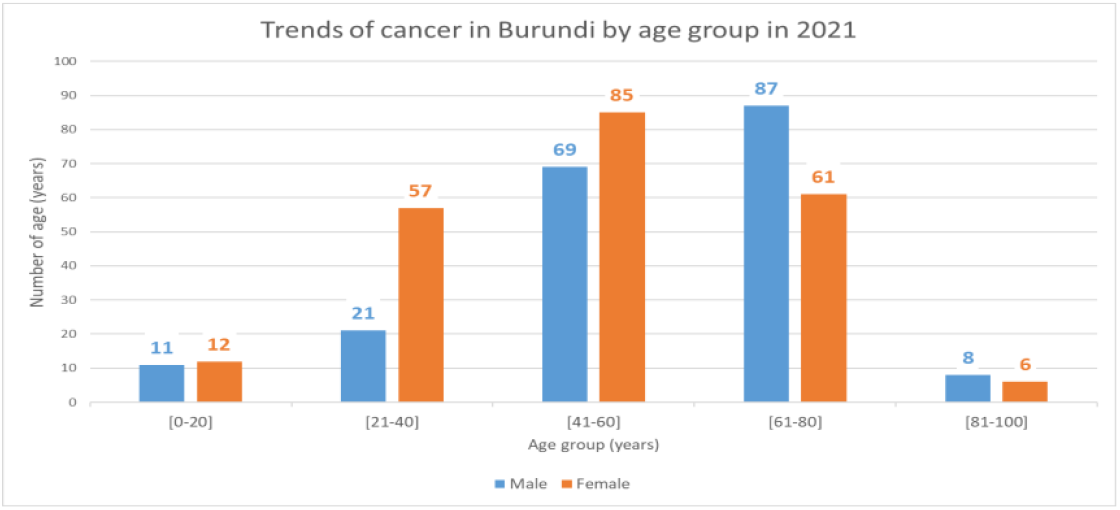
Trends of cancer cases by age groups for men and women.

### S3: Correlation matrix for between yearly data on diabetes, hypertension, cholesterol, BMI, diastolic blood pressure, systolic blood pressure in men and women population in Burundi between 1990 and 2014

**Figure.**
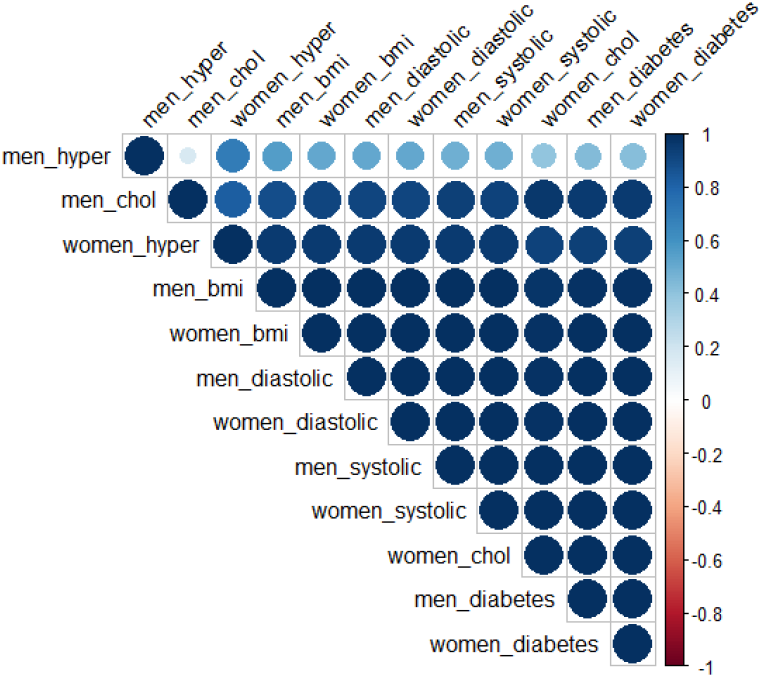
Correlation analysis between diabetes, hypertension, cholesterol, BMI, diastolic blood pressure, systolic blood pressure in men and women population.

